# Genome-wide association study of multiple neuropathology endophenotypes identifies novel risk loci and provides insights into known Alzheimer’s risk loci

**DOI:** 10.1101/2022.12.13.22282475

**Authors:** Lincoln M.P. Shade, Yuriko Katsumata, Steven A. Claas, Mark T. W. Ebbert, Erin L. Abner, Timothy J. Hohman, Shubhabrata Mukherjee, Kwangsik Nho, Andrew J. Saykin, David A. Bennett, Julie A. Schneider, The Alzheimer’s Disease Genetics Consortium, Peter T. Nelson, David W. Fardo, Alzheimer’s Disease Neuroimaging Initiative

**Affiliations:** Department of Biostatistics, College of Public Health, University of Kentucky, Lexington, KY; Sanders-Brown Center on Aging and Alzheimer’s Disease Research Center, University of Kentucky, Lexington, KY; Division of Biomedical Informatics, Department of Internal Medicine, University of Kentucky, Lexington, KY; Department of Neuroscience, University of Kentucky College of Medicine, Lexington, KY; Department of Epidemiology and Environmental Health, College of Public Health, University of Kentucky, Lexington, KY; Vanderbilt Memory & Alzheimer’s Center, Department of Neurology, Vanderbilt University Medical Center, Nashville, Tennessee; Department of Medicine, University of Washington, Seattle, WA; Department of Radiology & Imaging Sciences, Indiana University School of Medicine, Indianapolis, IN; Rush Alzheimer’s Disease Center and Department of Neurological Sciences, Rush Medical College, Chicago, IL; Rush Alzheimer’s Disease Center and Departments of Neurology and Pathology, Rush University Medical Center, Chicago, IL; Department of Pathology and Laboratory Medicine, University of Kentucky, Lexington, KY

**Keywords:** GWAS, Alzheimer’s, neuropathology, atherosclerosis, neurofibrillary tangles, cerebral amyloid angiopathy, TDP-43

## Abstract

**Background:** Alzheimer’s disease is highly heritable and exhibits neuropathological hallmarks of neurofibrillary tau tangles and neuritic amyloid plaques. Previous genome-wide association studies (GWAS) have identified over 70 genomic risk loci of clinically diagnosed Alzheimer’s disease. However, upon autopsy, many Alzheimer’s disease patients have multiple comorbid neuropathologies that may have independent or pleiotropic genomic risk factors. Autopsy data combined with GWAS provides the opportunity to study the genetic risk factors of individual neuropathologies.

**Methods:** We studied the genome-wide risk factors of eleven Alzheimer’s disease-related neuropathology endophenotypes. We used four sources of neuropathological data: National Alzheimer’s Coordinating Center, Religious Orders Study and Rush Memory and Aging Project, Adult Changes in Thought study, and Alzheimer’s Disease Neuroimaging Initiative. We used generalized linear mixed models to identify risk loci, followed by Bayesian colocalization analyses to identify potential functional mechanisms by which genetic loci influence neuropathology risk.

**Results:** We identified two novel loci associated with neuropathology: one *PIK3R5* locus (lead variant rs72807981) with neurofibrillary pathology, and one *COL4A1* locus (lead variant rs2000660) with cerebral atherosclerosis. We also confirmed associations between known Alzheimer’s genes and multiple neuropathology endophenotypes, including *APOE* (neurofibrillary tangles, neuritic plaques, diffuse plaques, cerebral amyloid angiopathy, and TDP-43 pathology); *BIN1* (neurofibrillary tangles and neuritic plaques); and *TMEM106B* (TDP-43 pathology and hippocampal sclerosis). After adjusting for *APOE* genotype, we identified a locus near *APOC2* (lead variant rs4803778) associated with cerebral amyloid angiopathy that influences DNA methylation at nearby CpG sites in the cerebral cortex.

**Conclusions:** rs2000660 is in strong linkage disequilibrium with a synonymous coding variant (rs650724) of *COL4A1*, providing a candidate functional variant. Two CpG sites affected by the cerebral amyloid angiopathy-associated *APOC2* locus were previously associated with dementia in an independent cohort, suggesting that the effect of this locus on disease may be mediated by DNA methylation. *BIN1* is associated with neurofibrillary tangles and neuritic plaques but not with amyloid pathology. *TMEM106B* is associated with hippocampal sclerosis and TDP-43 pathology but not the canonical Alzheimer’s disease pathologies. These findings provide insights into known Alzheimer’s disease risk loci by refining the pathways affected by these risk genes.

## Background

Aged individuals often accumulate multiple brain pathologies that contribute individually and synergistically to cognitive decline and dementia. Alzheimer’s disease (AD) is the most commonly diagnosed form of dementia and poses an enormous burden on human health and well-being. In the United States alone, over six million individuals are living with AD, and the disease imposes financial costs of treatment and care of over $300 billion annually [1].

The most common form of AD, late-onset AD (LOAD), presents in individuals aged 65 years and older and is highly heritable, with twin and family studies estimating its heritability at ∼60% [2–4]. Genome-wide association studies (GWAS) have to date identified over 70 genetic risk loci for LOAD that are involved in biological processes including amyloid precursor protein processing, immune response, and extracellular matrix regulation [5–7]. Most dementia GWAS to date have focused on clinically diagnosed LOAD or proxy outcomes based on family history of dementia [5–8]; however, late-onset dementia diagnosed as LOAD is increasingly recognized as a heterogeneous clinical syndrome that can reflect multiple underlying independent or co-occurring pathological processes that may have independent or pleiotropic genetic risk factors [9–11].

While neuritic plaques and neurofibrillary tangles (NFT) are present in the brains of the majority of people diagnosed with clinical AD, many people either do not have substantial AD neuropathology or have both AD and comorbid non-AD pathologies [11, 12]. For example, TDP-43 pathology contributes to an amnestic syndrome, limbic-predominant age-related TDP-43 encephalopathy (LATE), that presents similarly to LOAD [13]. However, LATE and LOAD exhibit differing patterns of pathogenesis in aged autopsy cohorts. LOAD pathology peaks in prevalence in individuals who die at age 95 years and decreases thereafter [14]. In contrast, prevalence of LATE neuropathologic change (LATE-NC) increases monotonically with age of death, with TDP-43 pathology often first appearing in the limbic region or entorhinal cortex and progressing to the neocortex [13, 14]. Hippocampal sclerosis (HS) of aging, a pathology characterized by uni-or bilateral neuronal death, gliosis, and atrophy of the hippocampus beyond normal ranges based on levels of AD pathology, commonly co-occurs with LATE-NC and is associated with severe cognitive impairment [13, 15].

In addition to proteinopathies, non-stroke vascular pathologies are common in elderly autopsied research participants and contribute to cognitive decline and dementia [16]. Cerebral amyloid angiopathy (CAA) is one such vascular pathology characterized by amyloid-beta deposition in cerebral blood vessels [17]. CAA often co-occurs with LOAD, but can independently contribute to cerebral injury by causing hemorrhages in the brain parenchyma [16, 17]. Infarcts of both small but grossly visible lacunar arteries and microscopically examined vessels (the latter referred to as microinfarcts) are also common in aged individuals and contribute to cognitive decline [16, 18]. Cerebral atherosclerosis and sclerosis of small cerebral blood vessels, called brain arteriolosclerosis, predispose individuals to infarcts and hippocampal sclerosis [19, 20]; moreover, they contribute to cognitive decline even after adjusting for the presence of related pathologies [21, 22]. Collectively, these factors reveal an increasingly complex and synergistic web of pathologies contributing to cognitive impairment and dementia, and demonstrate the need for continued study into genomic risk factors of dementia beyond those of clinically diagnosed LOAD.

The investigation of genetic risk of neuropathologies is an attractive approach for uncovering biological pathways involved in the development of LOAD and related dementias. Neuropathologies are commonly operationalized in aged autopsy cohorts as binary or semi-quantitative (or ordinal) neuropathology endophenotypes (NPE). Prior GWAS of neuropathology endophenotypes have confirmed known LOAD risk loci and have identified potential neuropathology risk loci [23–26]. However, continued recruitment in ongoing autopsy cohorts and methodological development for GWAS and downstream functional analysis provides additional opportunity for in-depth investigation into multiple neuropathologies studied across multiple cohorts.

In this study, we perform GWAS on eleven neuropathology endophenotypes collected in four high-quality studies with autopsy and genotype data available. Endophenotypes studied include AD-associated pathologies; LATE-NC; hippocampal sclerosis; vascular NPE including cerebral amyloid angiopathy, gross infarcts, microinfarcts, atherosclerosis, and arteriolosclerosis; and Lewy body pathology. We also perform downstream functional analyses to explore potential biological functional mechanisms of identified risk loci and provide insight into the shared genomic risk of neuropathologies.

## Methods

### Participants

We used genotype and neuropathology data from four high-quality data sources: the National Alzheimer’s Coordinating Center (NACC), the Religious Orders Study and the Rush Memory and Aging Project (ROSMAP), the Adult Changes in Thought (ACT) study, and the AD Neuroimaging Initiative (ADNI). Some participants in ROSMAP and ADNI also had neuropathology and genotype data available in NACC. In these cases, records in the NACC were preferentially kept in order to maximize sample size of the initial analysis using only NACC participants (see Statistical Analyses subsection).

All study participants were deceased and the resulting data de-identified for University of Kentucky investigators, and we exclusively used archival samples. Therefore, our study does not fall under the definition of “Human Subjects Research” according to the University of Kentucky Institutional Review Board because of NIH Exemption #4 – “involving the collection/study of data or specimens if publicly available, or/or recorded such that subjects cannot be identified.”

The present study used NACC data from 35 National Institute on Aging-funded Alzheimer’s Disease Research Centers (ADRCs). Individual ADRCs use different recruitment strategies and perform autopsies on-site, but neuropathology data at each ADRC are collected using a standard form (https://files.alz.washington.edu/documentation/np11-form.pdf) and submitted to NACC where they are aggregated and anonymized. The NACC Neuropathology (NP) data set based on the first version of this form was originally implemented in 2001 [27], and this analysis uses data from then through the December 2021 freeze. Participants were excluded if they did not have autopsy data available or if they were noted in the NP data set to have at least one of 19 conditions that could potentially bias results. These conditions include brain tumors, severe head trauma, and fronto-temporal lobar degeneration. (See Supplementary Table S1 for full list of variables used for exclusion criteria.)

ROSMAP has been previously described and consists of harmonized data from two longitudinal cohort studies: The Religious Orders Study (ROS) and the Rush Memory and Aging Project (MAP) [28]. ROS and MAP were both approved by an Institutional Review Board of Rush University Medical Center. All participants signed an Anatomic Gift Act, as well as informed and repository consents. ROS began in 1994 and has recruited over 1500 Catholic priests, nuns, and brothers across the United States. MAP started in 1997 and has enrolled more than 2300 community members in the greater Chicago area of northeastern Illinois. The ROSMAP NP data used in this study were received from Rush University Medical Center in January 2020.

The ACT study began in 1994 and recruited residents in the greater Seattle area aged 65 years and older without dementia at time of enrollment [29]. The study has expanded to include three cohorts with continued enrollment using the original enrollment criteria and has a current total of 4,960 participants across all three cohorts. The ACT NP data used in this study were obtained from Kaiser Permanente in May 2021.

The ADNI (https://adni.loni.usc.edu) was launched in 2003 as a public-private research partnership, led by Principal Investigator Michael W. Weiner, MD. The primary goal of ADNI has been to test whether serial magnetic resonance imaging, positron emission tomography (PET), other biological markers, and clinical and neuropsychological assessment can be combined to measure the progression of mild cognitive impairment and early AD. A subset of ADNI participants undergo autopsy and receive neuropathological phenotyping. The ADNI NP data used in this study was downloaded from the ADNI website in October 2021.

### Genotype data and quality control

Genotype data for all cohorts underwent imputation using the Trans-’Omics for Precision Medicine (TOPMed) Imputation Server and the TOPMed reference panel [30]. NACC and ACT raw genotype data were obtained from the September 2020 freeze of the Alzheimer’s Disease Genetics Consortium (ADGC) and subsequently imputed, while pre-imputed ROSMAP and ADNI genotype data were received from collaborators in the Hohman lab at Vanderbilt University in March 2021 and December 2021, respectively.

Quality control on genotype data was performed separately for each data source. Quality control and inclusion/exclusion criteria closely followed that used in our previous brain arteriolosclerosis GWAS [24]. Briefly, we included only participants labeled as non-Hispanic white or similar (European, etc.) if available. After imputation, we first identified and subsequently removed duplicate samples using the KING software “--duplicate” option [31]. We then removed participants without autopsy data available by merging genotype sample identification data with neuropathology data sets and retaining only samples present in both data sets. We then iteratively removed genetic variants and participants until no variants were missing in more than 5% of participants and no participants were missing more than 5% of variants (however, average genotype coverage was 99.7%, and no variants or participants were actually removed during this process). We then excluded participants with unusually high or low (±3 standard deviations from mean) genetic heterogeneity, as measured by the PLINK 1.9 “--het” flag [32]. Finally, we merged participants with the 1000 Genomes Phase 3 (1000 Genomes) cohorts [33] and performed principal components analysis (PCA) on a subset of independent variants (measured by pairwise *r*^2^ < 0. 2). We excluded participants with substantial non-European ancestry, as determined by distance from the 1000 Genomes EUR superpopulation centroid using the first two principal components (PCs).

### Defining and harmonizing neuropathology endophenotypes for analysis

In total, we combined or harmonized 11 neuropathology endophenotypes for analysis across the four studies. We note that there are differences in the way that some neuropathological data were collected across studies, and our strategy for harmonizing was informed by practical considerations for maximizing available samples sizes given the available endophenotypes. Thus, several synthetic NPE were created by merging existing NPE within a cohort or by harmonizing categorical variables from one cohort and continuous variables from another. A detailed listing of variables harmonized across cohorts to construct neuropathology endophenotypes for analysis is available in Supplementary Table S3. We briefly describe our harmonization methods below.

Arteriolosclerosis, Braak NFT stage, CAA, atherosclerosis, Consortium to Establish a Registry for Alzheimer’s Disease (CERAD) score for neuritic plaques, microinfarcts, and gross infarcts had variables in each cohort with directly comparable coding definitions and were straightforwardly renamed and combined with minimal recoding. Analyzed arteriolosclerosis, CAA, atherosclerosis, and CERAD score variables each had four stages with the following labels: 0 (“none”), 1 (“mild”), 2 (“moderate”), and 3 (“severe”). Microinfarcts and gross infarcts were labeled either 0 (“absent”) or 1 (“present”). The Braak NFT Stage variable analyzed followed the staging criteria previously described in the literature and had seven levels, ranging from 0 (absent NFT) to six (diffuse NFT throughout cortex and large loss of neurons) [34].

LATE-NC was recorded differently in several of the data sets and was harmonized to a four-level outcome variable following the simplified staging of TDP-43 pathology outlined in Figure 3B of the 2019 LATE working group report [13]. The following levels were used in the analyzed LATE-NC variable: 0, indicating lack of recorded TDP-43 proteinopathy; 1, indicating TDP-43 deposits in the amygdala only; 2, indicating deposits in the hippocampus or entorhinal cortex; and 3, indicating deposits in the neocortex. TDP-43 data was not available in ACT. In ROSMAP, TDP-43 pathology is recorded as a single variable following the same staging detailed above. In NACC and ADNI, the presence of TDP-43 pathology in each region of the brain is recorded as a separate binary indicator variable. To collapse TDP-43 pathology to a single ordinal variable in these data sets, we assigned a value based on the presence of the “highest” region where TDP-43 was present (e.g. a participant with TDP-43 pathology in both the hippocampus and the neocortex would be assigned a value of 3). Participants were labeled as 0 if they met two conditions: (1) they had recorded TDP-43 data available for at least one of the brain regions used for staging and (2) TDP-43 pathology was noted as absent in all the regions for which they had data available.

Diffuse amyloid plaque pathology was recorded as a four-stage Thal phase of amyloid deposition in NACC, ACT, and ADNI with the following levels: 0 (“none”), 1 (“mild”), 2 (“moderate”), and 3 (“severe”). In ROSMAP, diffuse plaques were examined and quantified in five regions (midfrontal cortex, entorhinal cortex, inferior parietal cortex, and hippocampus), then scaled by each region’s standard deviation and averaged. To discretize this continuous variable in ROSMAP participants to a four-level variable, as recorded in the other data sets, we assigned participants a value of 0 (“none”) if their averaged score was equal to 0, 1 (“mild”) if their score was higher than 0 but ≤ 0.5, 2 (“moderate”) if their score was between 0.5 and one, and 3 (“severe”) if their score was above 1. These labels roughly corresponded to score quartiles in ROSMAP.

Hippocampal sclerosis is recorded as a binary indicator of the presence or absence of pathology in the NACC NP data set form version 1-9, ROSMAP, and ACT. In versions 10 and 11 of the NACC NP form and in ADNI, HS pathology is recorded as being absent, unilateral, or bilateral. To harmonize, we dichotomized HS pathology as being present if either unilateral or bilateral pathology was indicated.

The Lewy body pathology variable we analyzed had four levels: 0, indicating absent Lewy body pathology in all regions examined or limited to the olfactory bulb; 1, indicating Lewy body pathology limited to the brainstem, including the substantia nigra; 2, indicating Lewy body pathology involving the limbic system or amygdala; and 3, indicating Lewy body involvement in the neocortex. In NACC and ROSMAP, Lewy body pathology was graded in ordinal variables with levels corresponding to the levels in the final harmonized outcome variable analyzed. In ADNI, a single variable was also used to record Lewy body pathology but included separate levels for (1) no pathology and olfactory-predominant and (2) limbic-predominant and amygdala-predominant pathology. These stages were collapsed into levels 0 and 2, respectively, for harmonization. In ACT, separate binary indicator variables were used to indicate presence of Lewy body pathology in each brain region checked. To harmonize, we created a new variable that coded Lewy body pathology stage according to the “highest” stage present in an individual (e.g. if pathology was present in both the amygdala and the neocortex, we assigned a value of 3).

### Statistical Analyses

#### Single-variant GWAS

We analyzed ordinal endophenotypes using proportional odds logistic mixed-effects models implemented in the POLMM R package [35]. We analyzed binary variables similarly with logistic mixed-effects models implemented in the SAIGE R package [36]. Covariates included age at death, sex, cohort, and the first 10 genetic PCs created using the PCA in Related Samples (PC-AiR) method in the GENESIS R package [37]. We included a dense genetic relationship matrix (GRM) as a random effect to account for relatedness between participants. An additive mode of inheritance was assumed in all analyses.

Analyses proceeded in two stages. In stage one, GRM were constructed using a pruned set of independent variants, defined as having a pairwise *r*^2^ < 0. 2 within moving windows of 15 kilobase pairs (kbp). Null models including fixed covariates and the GRM were then fitted using either the POLMM or SAIGE R packages. In stage 2, score tests were performed on each variant with a saddle-point approximation used to calculate p-values. We considered all variants with a p-value of *p* < 5 × 10^−8^ to be genome-wide significant. To identify independent risk loci, we clumped results using the “--clump” flag in PLINK 1.9 with the pairwise linkage-disequilibrium (LD) threshold set to *r*^2^ ≤ 0. 05 [32, 38].

We performed analyses both in individual cohorts separately and in a pooled data set with all participants. Single-variant analyses were first performed using NACC data. For all lead variants with a p-values 1 × 10^−6^, we attempted to validate the association in ROSMAP, ACT, and ADNI separately. Participants were then pooled into a single data set and genome-wide association analyses were performed again using all participants. As noted in the section on harmonization above, LATE-NC data was not available for ACT participants, so validation of LATE-NC-associated variants was not attempted in ACT.

#### Re-analysis of the APOE region

The region surrounding the *APOE* gene on Chromosome 19 is consistently the single strongest genetic risk factor for LOAD in GWAS. Three common forms of the *APOE* gene – ∈2, ∈3, and ∈4 – are present in our study populations, and the ∈2 and ∈4 alleles are associated with lower and higher risk of LOAD, respectively, relative to the ε3 allele. We therefore expected that variants in the *APOE* region, defined as the region within 200 kbp from the start and end transcription sites of *APOE*, would be associated with multiple NPE in our study. Moreover, we hypothesized that genetic variants in the *APOE* region may influence neuropathology risk independently of the effects of *APOE* ε alleles.

To test the hypothesis that genetic loci in the *APOE* region may affect neuropathology risk independently from *APOE* ε alleles, we re-analyzed variants in Chromosome 19 while adjusting for *APOE* ε genotype. We limited re-analysis to endophenotypes with at least one significant association signal within the *APOE* locus in the pooled single-variant association analysis described in the previous section. *APOE* genotypes were determined using the rs7412 and rs429358 variants according to the SNPedia online reference [39]. The ∈3/∈3 genotype was used as reference, and we included fixed-effect indicator variables to adjust for ∈2/∈2, ∈2/∈3, ∈3/∈4, and ∈4/∈4 genotypes. We chose this approach rather than adjusting for counts of ∈2 and ∈4 alleles because it is robust to potential non-linear effects of genotypes.

#### Gene-based analyses

Analyses that test for the aggregate effect of all variants in a gene region may increase power for detection of risk genes. We thus performed gene-based analyses for all neuropathology endophenotypes studied. Gene start and end sites were determined using GRCh38 gene regions. Variants were mapped to a gene if they were located within 10 kbp of the gene’s start or end sites. Using an unrelated set of the pooled participant data set, gene-based analyses were performed using MAGMA v1.10 [40]. The same fixed-effects covariates used in single-variant analyses were used, and the top variant PCs that accounted for 99.9% of variance in a gene’s region were used to test for significance using an F-test. We considered genes with *p* < 2. 5 × 10^−6^ to be significantly associated with endophenotypes.

#### Principal component outcome analyses

Many neuropathology endophenotypes are highly correlated with one another, and this phenomenon is not reflected in analyses using individual neuropathology endophenotypes as outcomes. Thus, we sought to perform genetic association analyses that could better account for the common co-occurrence of NPE. We first assessed the phenotype correlation of neuropathology endophenotypes used in the pooled GWAS using polychoric correlation and grouped endophenotypes by visually examining a dendrogram based on phenotype-phenotype correlations. Using the psych R package [41], for each group of endophenotypes, we then performed PCA on the matrix of neuropathology endophenotype outcomes in that group. The first PC (PC1) of each group was then used as the outcome variable for GWAS using SAIGE in a two-stage analysis. In the first stage, PC1 was regressed on fixed-effects covariates, with the GRM included as a random effect. Because the residuals of PC1 may not be normally distributed, the residuals of stage one were then rank inverse normalized and regressed on covariates and GRM again in stage two. Score tests were then performed on genetic variants and a saddle-point approximation used to estimate p-values.

#### Colocalization analyses

Colocalization analysis is a Bayesian analytical method that seeks to answer whether a single genetic locus drives genotypic-phenotypic associations in two or more traits. We used multiple sources of publicly available summary statistics from external studies as data sources for colocalization analyses. First, we downloaded Genotype-Tissue Expression Project (GTEx) v8 European ancestry quantitative trait loci (QTL) analysis summary statistics, which contains summary statistics for significant gene expression and splicing QTL variants (eQTL and sQTL, respectively) in 48 body tissues [42]. As an additional source of QTL data, we used gene expression and DNA methylation QTL (mQTL) analysis summary statistics from studies using tissue taken from the dorsolateral prefrontal cortex of ROSMAP participants [43]. These studies examined the associations of genetic variants with molecular traits and provide curated lists of significant QTL variants. Finally, we downloaded the summary statistics from a recent GWAS of LOAD for a targeted post hoc colocalization analysis in the *TMEM106B* and *GRN* genes [7].

For each neuropathology endophenotype outcome in our study, we first compiled a list of genetic variants with p-values ≤ 1 × 10^−5^ in the pooled single-variant genome-wide association analysis. We then queried the lists of significant QTL variants in GTEx and ROSMAP to identify neuropathology-associated QTL variants. For each genetic locus associated with neuropathology endophenotypes that had at least one significant QTL in either GTEx or ROSMAP, we performed colocalization analysis using the “coloc.abf” function in the coloc R package [44]. For ordinal variables, we chose dichotomizing cut points to determine case-control proportions. We used coloc’s default prior probability of colocalization (PrC) of *PrC* = 1 × 10^−5^ and considered a posterior *PrC* > 80% as a threshold for evidence of colocalization.

To investigate whether shared GWAS signals drive association among multiple neuropathology endophenotypes, we also performed colocalization analysis on loci with variants exceeding a p-value threshold of *p* < 1 × 10^−4^ and concordant effect direction for at least two NPE in the pooled single-variant analysis.

## Results

In total, 7,463 unique participants across all cohorts met our inclusion criteria and were included in our analyses; specifically, autopsy, and genotype data were available for these participants, and they passed our quality control measures for variant missingness, genetic heterogeneity, and ancestry. Included participants consisted of 5,625 participants from NACC; 1,183 from ROSMAP; 616 from ACT; and 39 from ADNI. Participant demographics are shown in Table 1. NACC participants died at a younger age (mean age at death 81 years) compared to ROSMAP (90 years) and ACT (88 years) participants, and had more balanced sex ratios, with 50% of NACC participants female vs. 68% and 58% in ROSMAP and ACT, respectively. NACC participants were also more likely to have an *APOE* ε4 allele (55%) vs. ROSMAP (25%) or ACT (28%). Across the 11 NPE harmonized across multiple cohorts, NACC and ADNI tended to show more advanced neuropathology than ROSMAP and ACT. Using CERAD neuritic plaque score as an example, 65% of NACC and 59% of ADNI participants were rated as “none” for CERAD score, whereas 33% of ROSMAP and 27% of ACT participants were rated as “severe.” This trend can be seen in most of the NPE harmonized across cohorts with the exceptions of CAA, LATE-NC, and microinfarcts (see Supplementary Table S2 for summary demographics for all neuropathology endophenotypes).

**Table 1:**
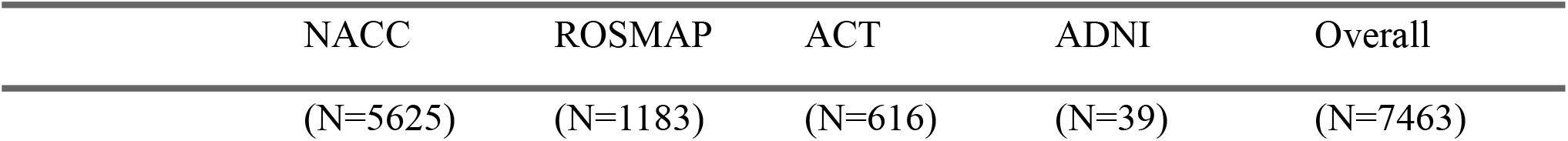
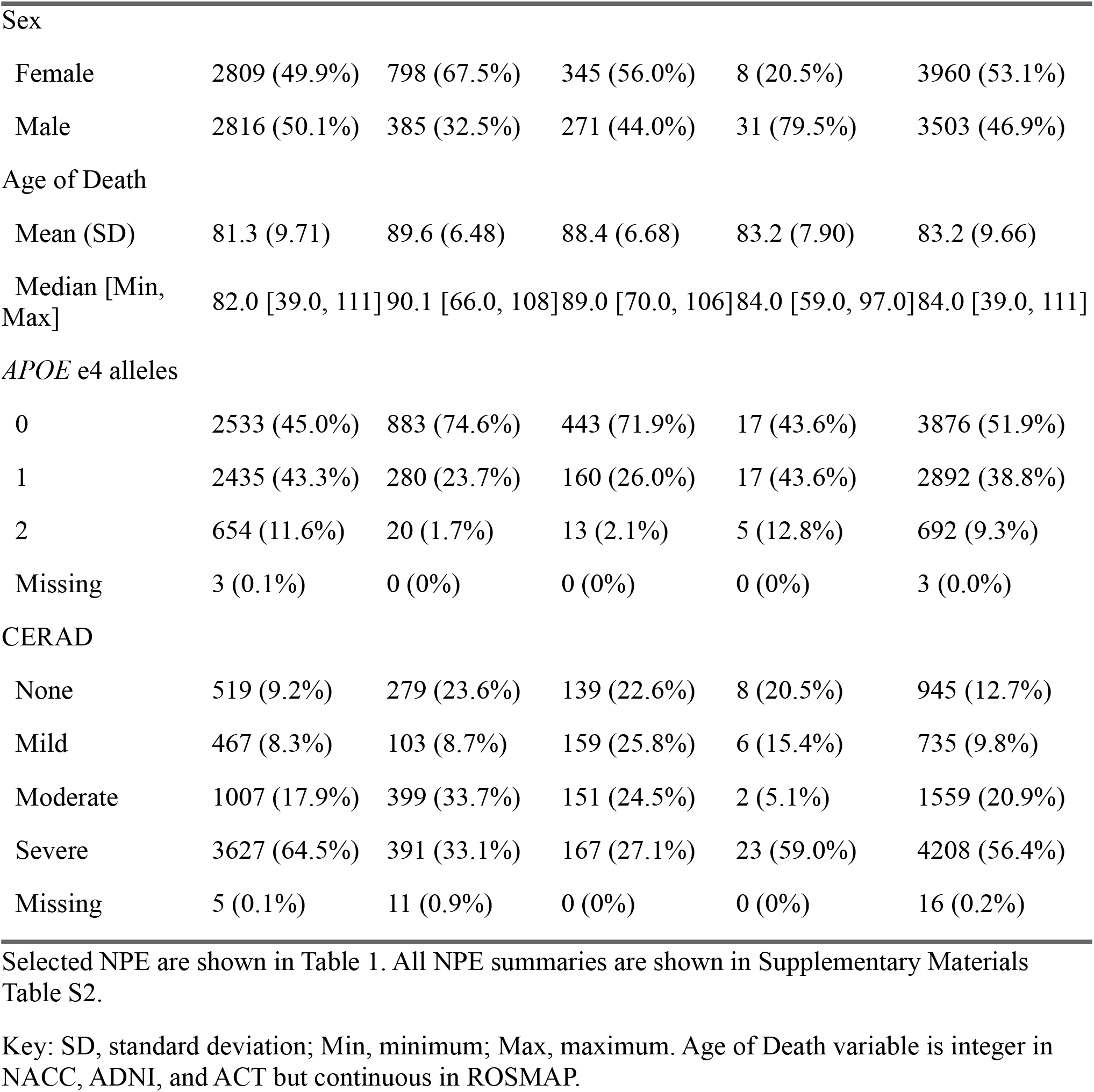
Participant Demographics

### Single-variant analysis identifies novel neuropathology-associated mapped to COL4A1 and PIK3R5

In the initial analysis using only NACC participants, variants within the *APOE* region were significantly associated with Braak NFT stage, CERAD score, CAA, and diffuse amyloid plaques. These associations were validated in all of the other data sets at a significance threshold of *p* ≤ 0. 05. No other loci were significantly associated with any neuropathology endophenotypes.

In total, 14 independent (*r*^2^< 0. 05) loci had lead variants that met the genome-wide significance threshold of *p* < 5 × 10^−8^ in the main pooled mega-analysis. Of these, four were duplicate signals in the *APOE* region, leaving 10 significant signals across 11 endophenotypes. Novel variant-endophenotype associations included (1) a variant 12 kbp upstream of *COL4A1*, rs2000660, associated with atherosclerosis in the circle of Willis (Figure 1) and (2) a *PIK3R5* intronic variant, rs72807981, associated with Braak NFT stage (Figure 2). Additionally, one locus with lead variant rs4803778 remained significantly associated with CAA in the *APOE* ε -adjusted analysis (Figure 3). Loci identified that have been previously associated with NPE or dementia included *APOE* with Braak stage, CAA, CERAD score, diffuse amyloid plaques, and LATE-NC; *BIN1* with Braak NFT stage; and *TMEM106B* with HS and LATE-NC (Table 2).

**Table 2:**
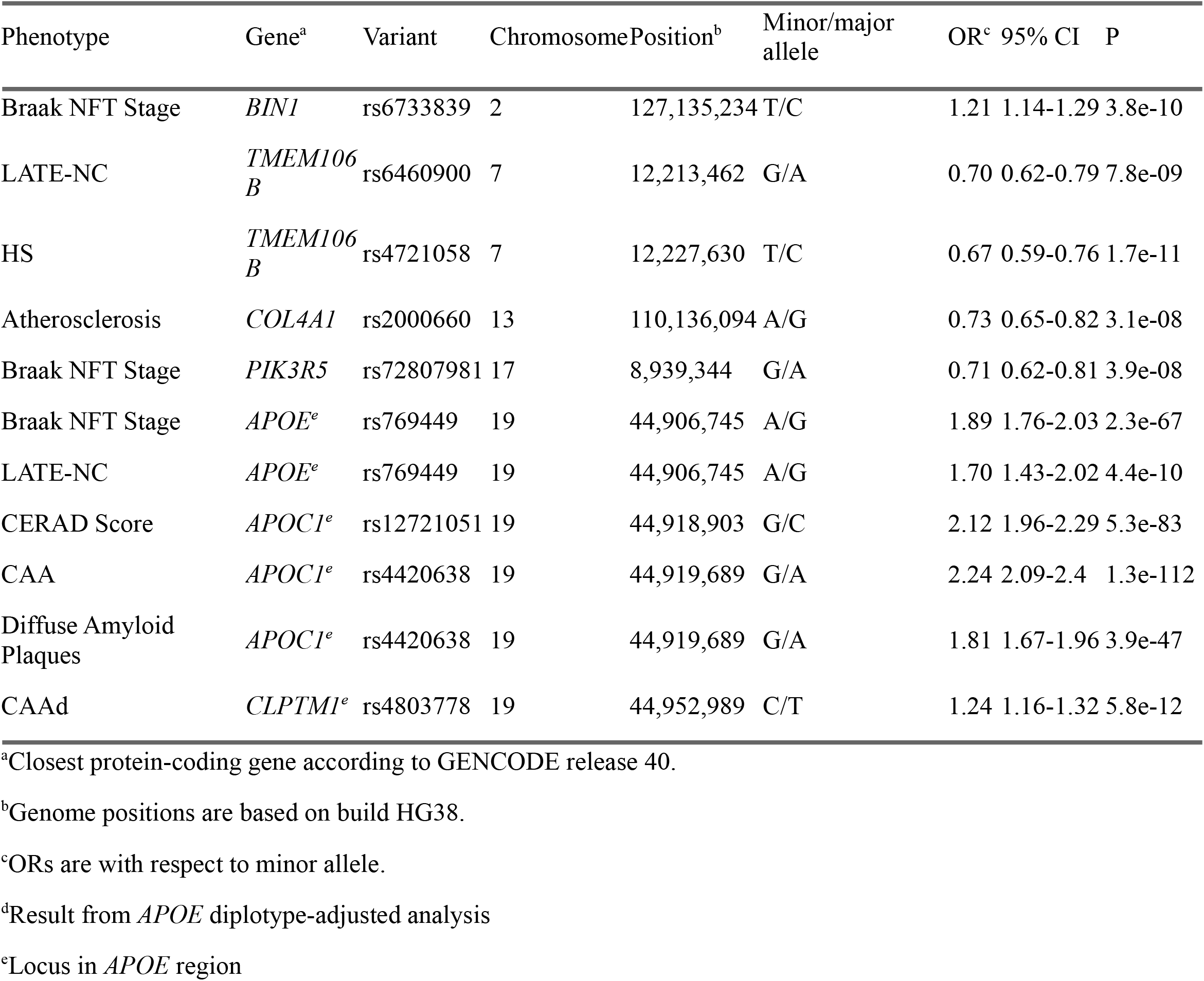
Significant NPE-Associated Loci in Mega-Analysis

| Phenotype | Gene<sup>a</sup> | Variant | Chromosome | Position<sup>b</sup> | Minor/major allele | OR<sup>c</sup> | 95% CI | P |
| --- | --- | --- | --- | --- | --- | --- | --- | --- |
| Braak NFT Stage | *BIN1* | rs6733839 | 2 | 127,135,234 | T/C | 1.21 | 1.14-1.29 | 3.8e-10 |
| LATE-NC | *TMEM106B* | rs6460900 | 7 | 12,213,462 | G/A | 0.70 | 0.62-0.79 | 7.8e-09 |
| HS | *TMEM106B* | rs4721058 | 7 | 12,227,630 | T/C | 0.67 | 0.59-0.76 | 1.7e-11 |
| Atherosclerosis | *COL4A1* | rs2000660 | 13 | 110,136,094 | A/G | 0.73 | 0.65-0.82 | 3.1e-08 |
| Braak NFT Stage | *PIK3R5* | rs72807981 | 17 | 8,939,344 | G/A | 0.71 | 0.62-0.81 | 3.9e-08 |
| Braak NFT Stage | *APOE<sup>e</sup>* | rs769449 | 19 | 44,906,745 | A/G | 1.89 | 1.76-2.03 | 2.3e-67 |
| LATE-NC | *APOE<sup>e</sup>* | rs769449 | 19 | 44,906,745 | A/G | 1.70 | 1.43-2.02 | 4.4e-10 |
| CERAD Score | *APOC1<sup>e</sup>* | rs12721051 | 19 | 44,918,903 | G/C | 2.12 | 1.96-2.29 | 5.3e-83 |
| CAA | *APOC1<sup>e</sup>* | rs4420638 | 19 | 44,919,689 | G/A | 2.24 | 2.09-2.4 | 1.3e-112 |
| Diffuse Amyloid Plaques | *APOC1<sup>e</sup>* | rs4420638 | 19 | 44,919,689 | G/A | 1.81 | 1.67-1.96 | 3.9e-47 |
| CAAd | *CLPTM1<sup>e</sup>* | rs4803778 | 19 | 44,952,989 | C/T | 1.24 | 1.16-1.32 | 5.8e-12 |

<sup>a</sup>Closest protein-coding gene according to GENCODE release 40.
<sup>b</sup>Genome positions are based on build HG38.
<sup>c</sup>ORs are with respect to minor allele.
<sup>d</sup>Result from *APOE* diplotype-adjusted analysis
<sup>e</sup>Locus in *APOE* region

**Figure 1:**
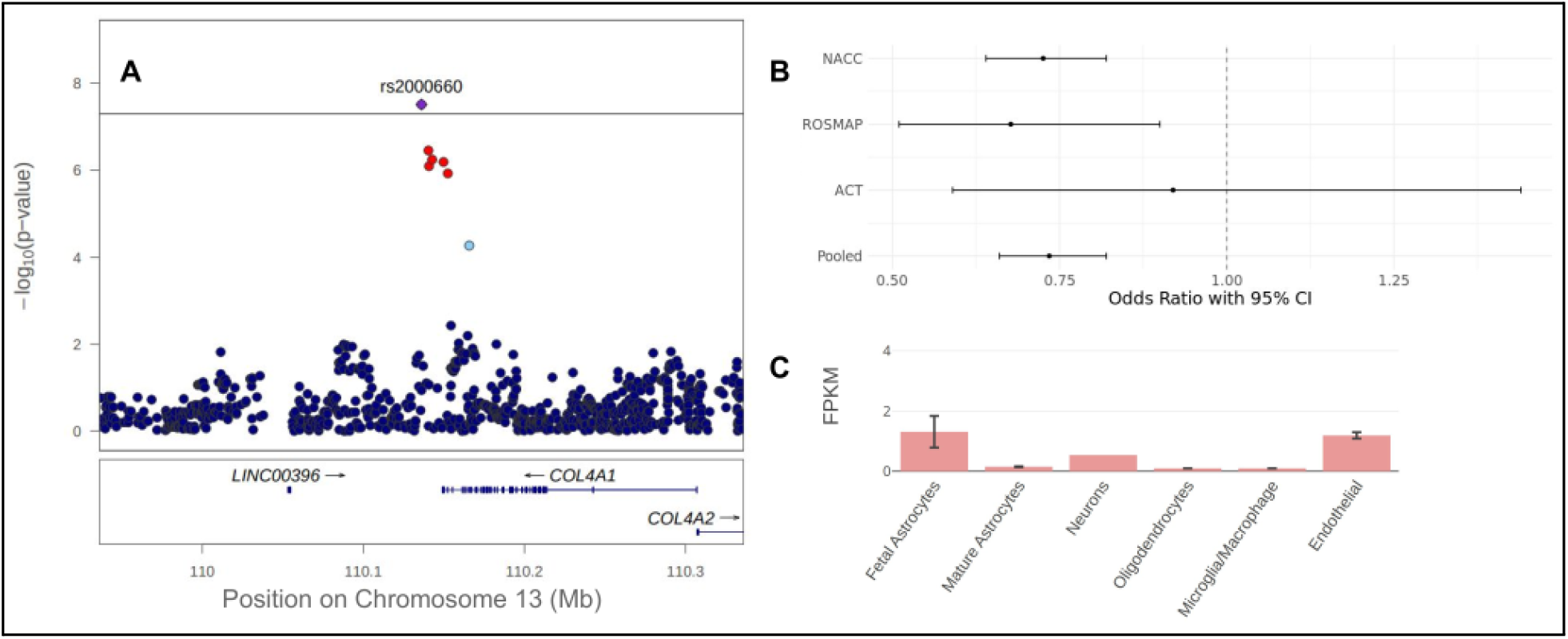
Locus near COL4A1 associates with cerebral atherosclerosis. A) Regional LocusZoom plot of associated locus [61]. Mb, megabase; uses hg19. B) Forest plot for individual and pooled cohorts for lead variant rs2000660 odds ratio and 95% confidence intervals. Lead variant reached a suggestive level of significance in NACC analysis (*OR* = 0. 72, *p* = 5. 8 × 10^−7^) and was nominally validated in ROSMAP (OR = 0.68, p = 0.0079). ADNI results are not shown due to wide confidence interval (OR = 1.25, 95% CI = 0.08-18.84). C) Human brain cell-type expression profile of *COL4A1* in Zang et al. (2016) [53]. *COL4A1* is preferentially expressed in fetal astrocytes and endothelial cells with lower expression in neurons. FPKM, Fragments Per Kilobase of transcript per Million mapped reads.

**Figure 2:**
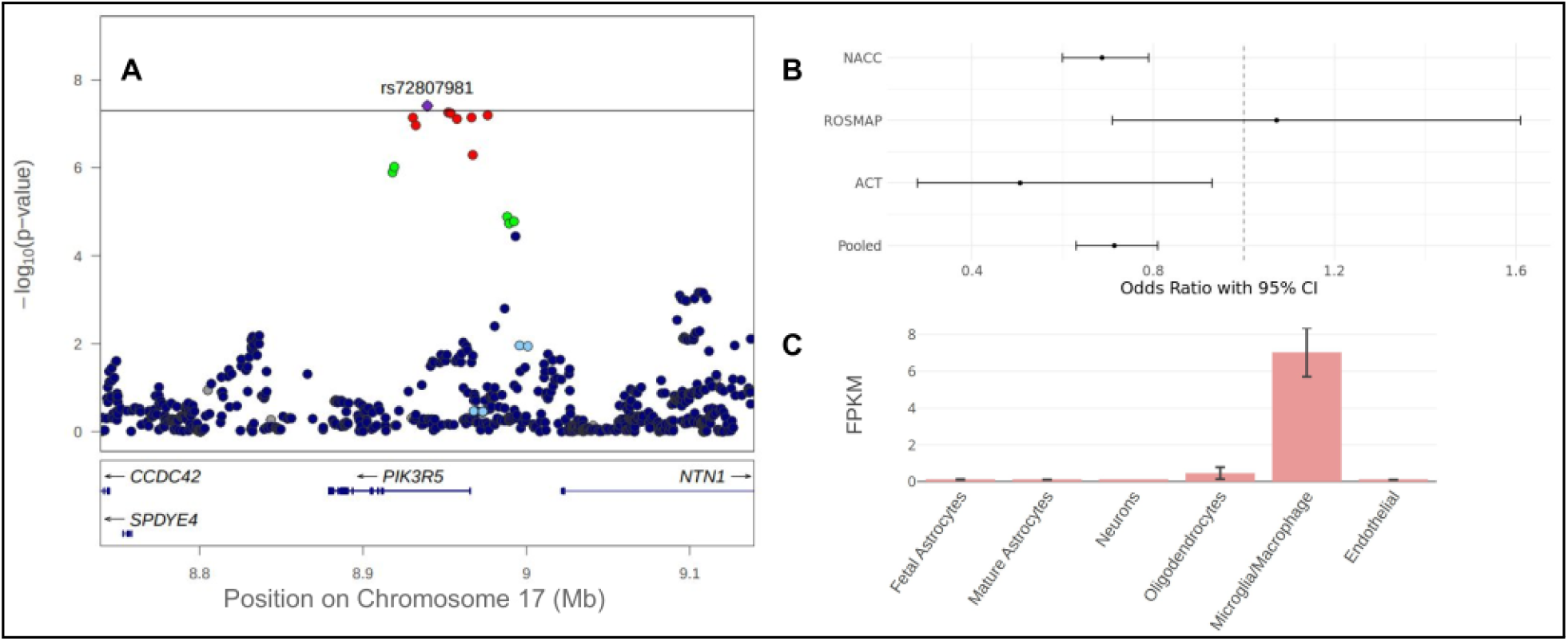
Intronic PIK3R5 locus associates with Braak stage. A) Regional LocusZoom plot of Braak-associated locus. Mb, megabase; uses hg19. B) Forest plot for individual and pooled cohorts for rs72807981 odds ratio and 95% confidence intervals. Lead variant rs72807981 reached a suggestive level of significance in NACC analysis (*OR* = 0. 69, *p* = 1. 8 × 10^−7^) and was nominally validated in ACT (*OR* = 0. 50, *p* = 0. 0028). C) Human brain cell-type expression profile of *PIK3R5* in Zang et al. (2016) [53]. *PIK3R5* is primarily expressed in microglia. FPKM, Fragments Per Kilobase of transcript per Million mapped reads.

**Figure 3:**
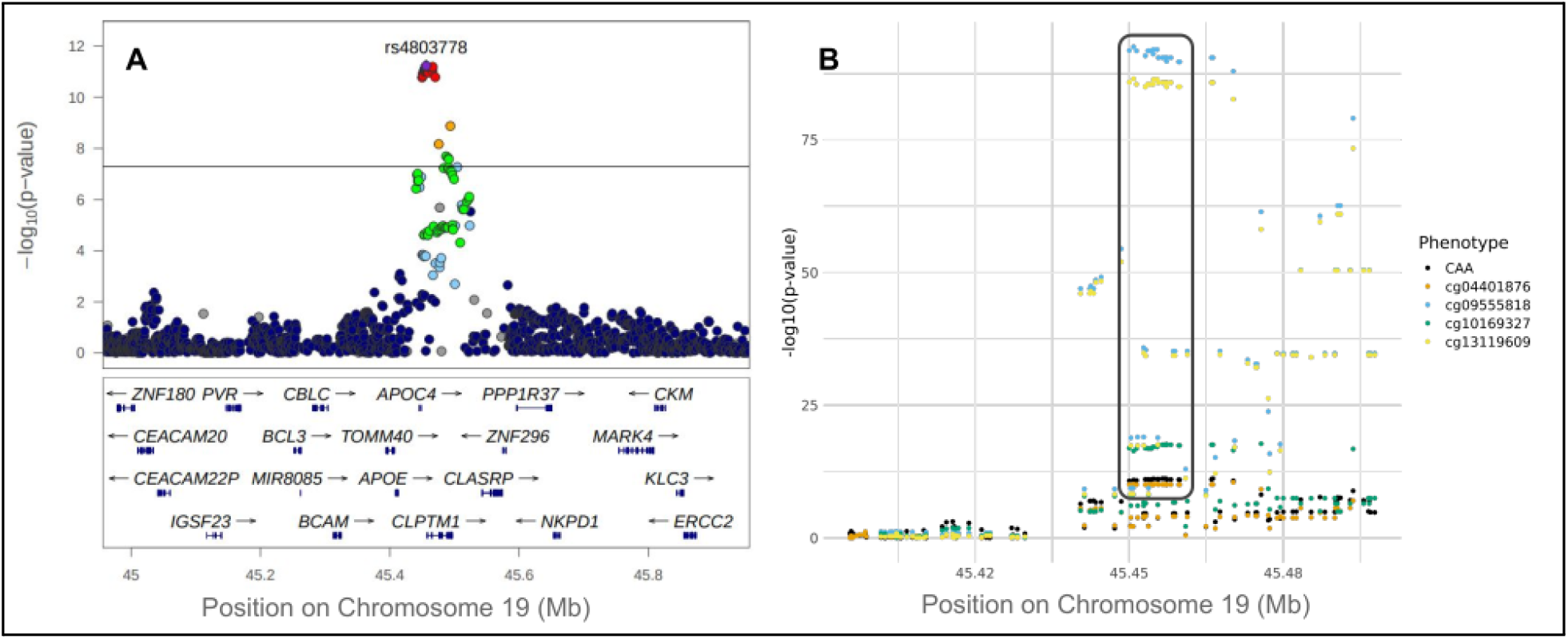
Locus in APOE region associates with CAA and colocalizes with mQTL in ROSMAP. A) Regional LocusZoom of associated locus. Mb, megabase; uses hg19. Eighteen genes are not shown due to space considerations. B) Regional plot showing CAA associations and colocalization with four mQTL in ROSMAP with CpG sites: cg04401876, cg09555818, cg10169327, and cg13119609. Posterior probability of colocalization equals 97% with each of the four mQTL. The darkly bordered box indicates the region with SNPs most associated with CAA and the four colocalizing mQTL.

### Gene-based analysis confirms association between genes in APOE region and multiple neuropathology endophenotypes

Gene-level analyses further corroborated our single-variant analyses, identifying gene associations with neuropathology endophenotypes in regions with significant variant-phenotype associations. *APOE* was significantly associated with NFT, diffuse plaques, CAA, neuritic plaques, and LATE-NC. CYP27C1, located approximately 50 kbp downstream of *BIN1*, was significantly associated with NFT. *TMEM106B* was associated with both HS and LATE-NC.

### Analysis of neuropathology endophenotypic correlations identifies Alzheimer’s-associated and vascular clusters

Visual inspection of a dendrogram (Supplementary Figure S1) of neuropathology endophenotypes based on polychoric correlations reveals two primary clusters of NPE: (1) a cluster of AD-or amyloid-associated endophenotypes consisting of Braak NFT stage, CERAD score, diffuse amyloid, and CAA; and (2) a cluster of vascular endophenotypes consisting of atherosclerosis, arteriolosclerosis, gross infarcts, and microinfarcts. PC1 in the AD cluster accounted for 66% of variance in the cluster, while PC1 in the vascular cluster accounted for 50% of the variance in that cluster. Only the *APOE* region was significantly associated with the AD cluster PC1, while no loci were significantly associated with the vascular PC1.

### Colocalization analysis identifies colocalization between CAA and DNA methylation in APOC2 region

Because CAA was the only NPE to have an associated locus in the *APOE* region with a signal independent of *APOE* ε diplotype, this region was excluded from endophenotype-endophenotype colocalization analysis. Pairs of endophenotypes with the same variant reaching a p-value threshold of *p* < 1 × 10^−5^ with concordant effect directions included (1) Braak NFT stage and CERAD score in the *BIN1* region on Chromosome 2 and (2) HS and LATE-NC in the *TMEM106B* region on Chromosome 7 (Figure 4). Both loci showed a high probability of colocalization, with both having *PrC* > 90%.

**Figure 4:**
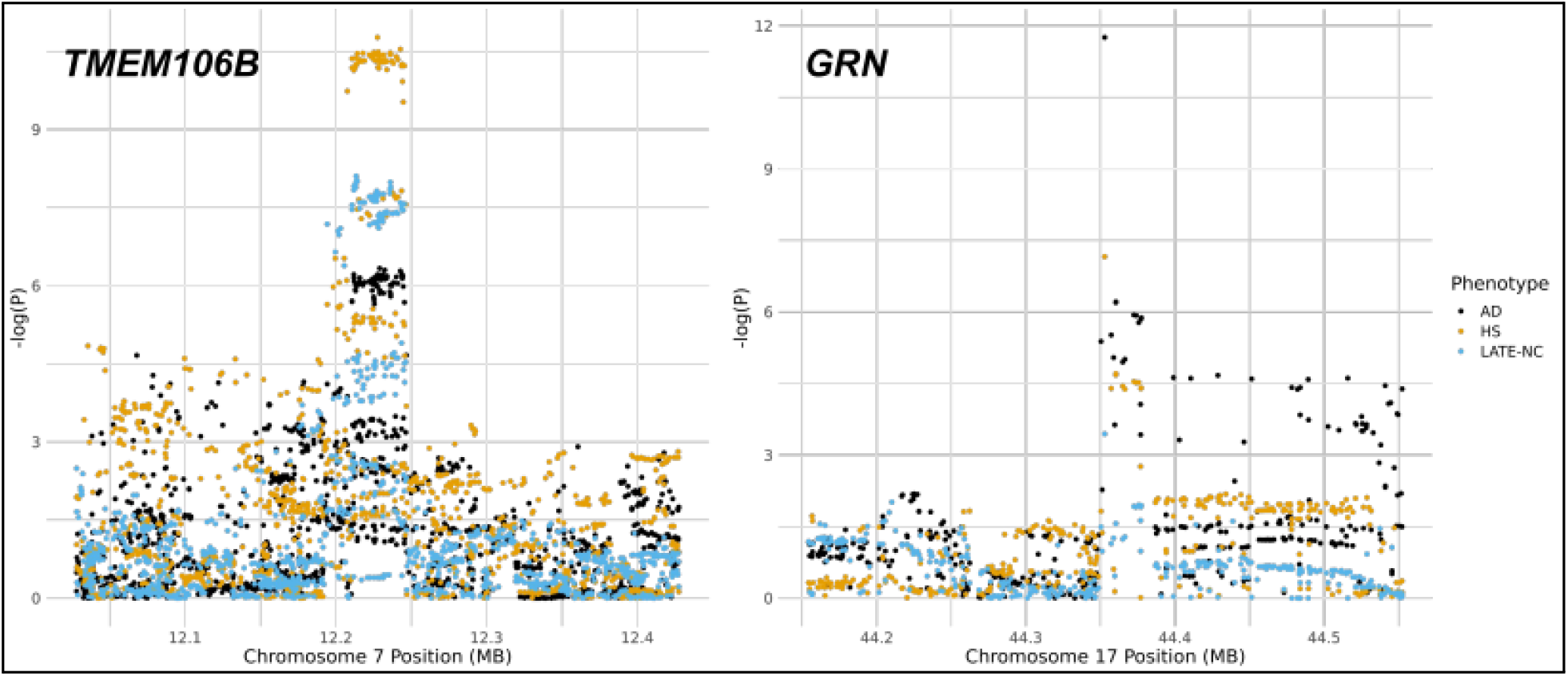
HS, LATE-NC, and AD colocalize at TMEM106B and GRN loci. Genome positions aligned to hg38. A) *TMEM106B* locus. Posterior probability of colocalization (PrC) equals 91% for each pair of phenotypes. B) GRN locus. PrC is greater than 99.9% between HS and AD. PrC equals 90% between LATE-NC and AD. PrC equals 89% between HS and LATE-NC. Key: MB, mega-basepairs; HS, hippocampal sclerosis; AD, clinical or proxy Alzheimer’s disease; LATE-NC, limbic-predominant age-related TDP-43 encephalopathy neuropathologic change.

Multiple suggestive NPE loci showed evidence of colocalization with eQTL in GTEx. A locus within *GRN* suggestively associated with HS (*p* = 6. 9 × 10^−8^) colocalized with *GRN* expression (*PrC* > 99. 9%), and the *TMEM106B* locus associated with HS and LATE-NC colocalized with *TMEM106B* eQTL. The CAA-associated *APOC2* locus independent of *APOE* ∈ diplotype also colocalized with *APOE* and *APOC2* eQTL in GTEx (*PrC* = 95%) and with four mQTL in ROSMAP (*PrC* = 97% for each of the four mQTL, see Figure 3).

## Discussion

We performed GWAS and downstream in silico functional analyses for eleven neuropathology endophenotypes across four high-quality neuropathology cohorts with a maximum sample size of 7,463 participants. Our work builds on previous genetic association studies of NPE, Alzheimer’s, and related dementias and provides another attempt to better understand the complex associations between different neuropathologies and genetic risk factors [7, 10, 23–25, 45]. Our study has several important limitations, including sample size and heterogeneity in study design. However, we discovered intriguing new loci mapped to *COL4A1* and *PIK3R5* associated with atherosclerosis in the circle of Willis and Braak stage for neurofibrillary tangles, respectively. We also investigated known loci in *BIN1, APOE*, and TMEM106B to provide additional context on their associations with multiple NPE.

### Limitations

There are limitations to the approach we took in this study. The four data sources we used, while all of high quality, used different study designs and recruitment strategies. NACC participant data arises from recruitment at each ADRC and likely consists of a more clinical population, whereas MAP and ACT participants may be more representative of the broader aged communities from which they are recruited. ROS participants include Catholic brothers, sisters, and priests and may represent a population more highly educated than the United States as a whole. These differences are reflected in the relative frequencies and distributions of NPE between cohorts, with NACC and ADNI tending to show more severe pathology than ACT and ROSMAP, and their mean ages of death were markedly lower (Tables 1 and S2). Additionally, there is likely substantial heterogeneity in the way that neuropathology data were collected and graded both within and between studies. We review these potential limitations briefly in our previous GWAS of brain arteriolosclerosis [24].

Additionally, while recruitment, autopsy, and genotyping of the cohorts used is ongoing, the sample sizes available for genome-wide investigation of NPE genetic risk is small relative to their clinical counterparts. A recent GWAS of AD included over 111,000 clinically diagnosed or “proxy” (based on family history) AD cases and nearly 700,000 controls. In contrast, our largest analyzed sample size for CERAD score totaled 7,447 participants, while LATE-NC totaled only 2,224. These samples reflect immense progress in autopsy cohorts from even eight years ago and will only continue to increase, but available sample sizes still pose limitations on power of GWAS and similar ‘omics approaches to studying disease risk using neuropathology endophenotypes. A strength of our study is that data on multiple neuropathology is available for each participant, allowing for multivariate approaches to studying NPE. However, as our PCA-based multivariate analyses did not identify any risk loci not identified in single-outcome analyses, this approach alone is not sufficient overcome sample size limitations.

### COL4A1 and atherosclerosis

One locus on Chromosome 12q34 with lead variant rs2000660 (minor allele frequency [*MAF*] = 9. 9% in pooled data set) located 12 kbp upstream of *COL4A1* was significantly associated with atherosclerosis in the circle of Willis (odds ratio [*OR*) = 0. 71, *p* = 3. 9 × 10^−8^). *COL4A1* in the brain is preferentially expressed in endothelial cells (Figure 1) and codes for a component of collagen IV, an important component of basal lamina. In previous GWAS, researchers have reported the *COL4A1*/COL4A2 locus to be associated with numerous other vascular phenotypes, including peripheral artery disease, coronary artery disease, stroke, and arteriolar stiffness [48]. The *COL4A1*/COL4A2 locus has also been found to be associated with rare familial cerebrovascular diseases and lacunar ischemic stroke[49, 50]. In a recent GWAS, rs2000660 in particular was a risk variant for migraines [51]. The relevance of the *COL4A1* locus to cerebral vascular traits is thus highly supported by previous research, and the biological role of collagen IV to vascular disease is possibly related to disruption of the extracellular matrix [48].

rs2000660 was not a QTL in GTEx or ROSMAP. However, rs650724, a variant in high LD with rs2000660 (*r*^2^ = 0. 84), is a synonymous coding variant (p.Ser1600Ser in ENST00000375820.10; p.Ser319Ser in ENST00000650424.1) for *COL4A1*. rs2000660 was not nominally associated with any other vascular NPE in our study, and a previous GWAS of cerebral atherosclerosis using ROSMAP participants did not identify the *COL4A1* as a risk locus [25]. However, the sample size of the previous study was significantly smaller than the one used in the present study (1,325 vs 6,959). Indeed, in the present study, rs2000660 reached genome-wide significance only in the pooled mega-analysis, though it was nominally significant in the ROSMAP-only validation analysis (*p* = 0. 0079).

### PIK3R5 and Braak NFT stage

The other identified novel genome-wide significant locus located on Chromosome 17p13 was associated with Braak NFT stage and had a lead variant of rs72807981 (*OR* = 0. 71, *p* = 3. 9 × 10^−8^, *MAF* = 6. 3%), an intronic variant within *PIK3R5*, which codes for a phosphatidylinositol 3-kinase involved in cell growth, motility, and survival. This variant was suggestively associated with Braak NFT stage in NACC (*OR* = 0. 69, *p* = 1. 8 × 10^−7^) and was nominally validated in ACT (*OR* = 0. 50, *p* = 0. 028). The same variant was also nominally associated with neuritic plaques in the pooled analysis (*OR* = 0. 82, *p* = 0. 0047) but was not nominally associated with any other AD NPE. rs72807981 is not a QTL in GTEx or ROSMAP, nor were any of the variants in high LD with it in our study. There is previous research suggesting that *PIK3R5* may be involved in the development of NFT. One study compared differential mRNA expression in aged adults with Braak NFT stage VI vs. non-demented controls and found *PIK3R5* to be more highly expressed in cases than controls (false-discovery rate [*FDR*] = 0. 01) [52]. Interestingly, *PIK3R5* is expressed preferentially in microglial cells in humans (see Figure 2), suggesting that its association with neurofibrillary pathology may be immune-mediated [53].

### APOE and NPE

Multiple variants in the *APOE* region were associated with pathognomonic AD NPE, including Braak stage, CERAD score, diffuse amyloid plaques, and CAA. Variants in the *APOE* region were also associated with LATE-NC, which is consistent with previous genetic association studies of NPE [23, 45]. While multiple loci in this region were significantly associated with each of these four NPE, only one CAA-associated locus with lead variant rs4803778 remained significant (*OR* = 1. 24, *p* = 5. 8 × 10^−12^) when adjusting for *APOE* ∈ diplotype. rs4803778 was in high LD (*r*^2^ = 0. 97) with rs4803779, which was also significantly associated with CAA (*OR* = 1. 24, *p* = 6. 7 × 10^−12^), is a lead mQTL in ROSMAP for five methylation sites (*p* = 3. 3 × 10^−91^ for top site cg09555818; see Figure 3), and colocalizes with four of these mQTL (*PrC* = 97% for all four sites). Furthermore, two of these associated methylation sites, including cg09555818, were significantly associated with dementia in the Generation Scotland: Scottish Family Health Study [54]. These results indicate that this locus may be associated with CAA through epigenetic regulation. However, this locus is located approximately 30 kbp downstream of the *APOE* coding end site and is situated between the *APOC4, APOC2*, and *CLPTM1*. While *APOE* may be the most likely target gene of this locus due to its known association with CAA, this inference is not straightforward. In GTEx, this locus is a stronger eQTL of APOC2 than *APOE*, although it is strongly significant and colocalizes with expression of both genes in multiple tissues. Future work should examine the regulatory role of the identified methylation sites and test them for association with CAA and other AD NPE.

### HS and LATE-NC

One intronic locus of *TMEM106B* was significantly associated with both HS (*OR* = 1. 50, *p* = 1. 5 × 10^−11^) and LATE-NC (*OR* = 1. 50, *p* = 1. 5 × 10^−11^). Additionally, a locus within *GRN* was suggestively associated with HS (*OR* = 0. 72, *p* = 6. 9 × 10^−8^). Both of these genes have been found previously to be associated with frontotemporal lobar degeneration [55, 56], and in a recent GWAS by Bellenguez et al., both of these genes were identified as risk factors for LOAD [57]. We downloaded available summary statistics from the GWAS Catalog and performed additional colocalization analyses at these loci to investigate if these loci were shared between clinical LOAD phenotypes and NPE [58]. We found that HS (*PrC* = 91%) and LATE-NC (*PrC* = 91%) colocalized with AD at both the *TMEM106B* (HS and LATE-NC both *PrC* = 91%) and *GRN* (HS *PrC* > 99. 9%, LATE-NC *PrC* = 90%) loci (Figure 4). HS and LATE-NC also colocalized with each other at these loci, as discussed above. These results indicate that HS, LATE-NC, and AD likely share causal loci at these genes.

### BIN1, neurofibrillary tangles, and neuritic plaques

A locus approximately 30 kbp downstream of *BIN1* on Chromosome 2q14 was significantly associated with Braak NFT stage (*OR* = 1. 21, *p* = 3. 8 × 10^−10^) and suggestively associated with CERAD score for neuritic plaques (*OR* = 1. 20, *p* = 6. 6 × 10^−8^). In our colocalization analysis, we verified that the same locus drives association signals with Braak NFT stage and CERAD score. In prior GWAS, this locus is second only to *APOE* for strength of association with LOAD [7]. However, the lead variant in this locus, rs6733839, was associated with neither diffuse amyloid plaques (*OR* = 1. 05, *p* = 0. 15) nor CAA (*OR* = 1. 02, *p* = 0. 49). Previous research supports the hypothesis that *BIN1* is associated with LOAD through its effect on NFT rather than amyloid pathology. In one study, researchers identified that *BIN1* variant rs744373 is associated with tau-PET but not amyloid-PET levels [59]. In another study, authors found that *BIN1* protein expression is correlated with NFT pathology but not with diffuse or neuritic plaques [60]. As neuritic plaques contain dying nerve cell processes with aberrant tau fibrils identical to those seen in neurofibrillary tangles [12], our findings are also consistent with the hypothesis that *BIN1* influences AD risk primarily through tau rather than amyloid pathogenic processes.

## Conclusions

In conclusion, we identified several promising novel loci associated with NPE and validated multiple known risk loci for AD using NPE. We also provided additional context and consideration for relationships between specific risk loci and different NPE. Overall, our study provides additional potential avenues of investigation into the relationship between genomics, Alzheimer’s disease, and neuropathology.

## Data Availability

All code used for data preparation and analysis is available at https://www.github.com/lincoln-shade/np_phewas. ROSMAP data can be requested at https://www.radc.rush.edu and https://www.synapse.org. ADGC data is can be requested from NIAGADS at https://www.niagads.org/resources/related-projects/alzheimers-disease-genetics-consortium-adgc-collection. NACC neuropathology data can be requested at https://naccdata.org/. ACT data can be requested at https://actagingresearch.org/. ADNI data can be downloaded at https://adni.loni.usc.edu/.

## List of abbreviations

AD: Alzheimer’s disease;
LOAD: late-onset Alzheimer’s disease;
GWAS: genome-wide association study;
NFT: neurofibrillary tangle;
LATE: limbic-predominant age-related TDP-43 encephalopathy;
LATE-NC: LATE neuropathologic change;
HS: hippocampal sclerosis;
CAA: cerebral amyloid angiopathy;
NPE: neuropathology endophenotype;
NACC: National Alzheimer’s Coordinating Center;
ROSMAP: the Religious Orders Study and the Rush Memory and Aging Project;
ACT: the Adult Change in Thought Study;
ADNI: AD Neuroimaging Initiative;
ADRC: Alzheimer’s Disease Research Center;
NP: neuropathology;
ROS: Religious Orders Study;
MAP: Rush Memory and Aging Project;
PET: positron emission tomography;
TOPMed: Trans-’Omics for Precision Medicine;
ADGC: Alzheimer’s Disease Genetics Consortium;
PCA: principal components analysis;
GRM: genetic relationship matrix;
PC-AiR: PCA in Related Samples;
kbp: kilobase pairs;
LD: linkage disequilibrium;
PC1: first principal component;
GTEx: Genotype-Tissue Expression Project;
QTL: quantitative trait locus;
eQTL: expression QTL;
sQTL: splicing QTL;
mQTL: methylation QTL;
OR: odds ratio;
PrC: probability of colocalization.

## Declarations

### Ethics approval and consent to participate

All study participants were deceased and the resulting data de-identified, and we exclusively used archival samples. Therefore, our study does not fall under the definition of “Human Subjects Research” according to the University of Kentucky Institutional Review Board because of NIH Exemption #4 – “involving the collection/study of data or specimens if publicly available, or/or recorded such that subjects cannot be identified.”

### Availability of data and materials

All code used for data preparation and analysis will be available at https://www.github.com/lincoln-shade/np_phewas upon publication. ROSMAP data can be requested at https://www.radc.rush.edu and https://www.synapse.org. ADGC data can be requested from NIAGADS at https://www.niagads.org/resources/related-projects/alzheimers-disease-genetics-consortium-adgc-collection. NACC neuropathology data can be requested at https://naccdata.org/. ACT data can be requested at https://actagingresearch.org/. ADNI data can be downloaded at https://adni.loni.usc.edu/.

### Competing interests

J.A.S. reported personal fees from Observational Study Monitoring Board Framingham, Observational Study Monitoring Board Discovery (National Institute of Neurological Disorders and Stroke), and Takeda Pharma. A.J.S. reported support from Avid Radiopharmaceuticals, a subsidiary of Eli Lilly (in kind contribution of PET tracer precursor); Bayer Oncology (Scientific Advisory Board); Eisai (Scientific Advisory Board); Siemens Medical Solutions USA, Inc. (Dementia Advisory Board); NIH NHLBI (MESA Observational Study Monitoring Board); and Springer-Nature Publishing (Editorial Office Support as Editor-in-Chief, Brain Imaging and Behavior). All other authors declare that they have no competing interests.

### Funding

The author(s) disclosed receipt of the following financial support for the research, authorship, and/or publication of this article: R56AG057191; F30NS124136; P30AG028383; the University of Kentucky Center for Clinical and Translational Science TL-1 Fellowship [grant number TL1TR001997]; the National Center for Advancing Translational Sciences [grant number UL1TR001998]; and the Dean of the College of Medicine, University of Kentucky. The content is solely the responsibility of the authors and does not necessarily represent the official views of the NIH or the University of Kentucky.

Genotyping was supported by the Alzheimer’s Disease Genetics Consortium through the National Institute of Aging [grant numbers U01 AG032984, RC2AG036528].

Samples from the National Cell Repository for Alzheimer’s Disease (NCRAD), which receives government support under a cooperative agreement grant (U24 AG21886) awarded by the National Institute on Aging (NIA), were used in this study. We thank contributors who collected samples used in this study, as well as patients and their families, whose help and participation made this work possible; Data for this study were prepared, archived, and distributed by the National Institute on Aging Alzheimer’s Disease Data Storage Site (NIAGADS) at the University of Pennsylvania (U24-AG041689-01).

We thank the study participants and staff of the Rush Alzheimer’s Disease Center. The Religious Orders Study and the Rush Memory and Aging Project are supported by grants from the National Institutes of Health: [grant number P30AG10161, P30AG72975, R01AG15819, R01AG17917, R01AG22018, R01AG33678, R01AG34374, R01AG36042, R01AG40039, R01AG042210, U01AG46152, U01AG61356, R01AG47976, R01AG43379, RF1AG54057, R01AG56352, R01NS78009, and UH2NS100599], and the Illinois Department of Public Health.

The Adult Changes in Thought Study is funded through the National Institute on Aging [grant number U19AG066567].

Data collection and sharing for this project was funded by the Alzheimer’s Disease Neuroimaging Initiative (ADNI) (National Institutes of Health Grant U01 AG024904) and DOD ADNI (Department of Defense award number W81XWH-12-2-0012). ADNI is funded by the National Institute on Aging, the National Institute of Biomedical Imaging and Bioengineering, and through generous contributions from the following: AbbVie, Alzheimer’s Association; Alzheimer’s Drug Discovery Foundation; Araclon Biotech; BioClinica, Inc.; Biogen; Bristol-Myers Squibb Company; CereSpir, Inc.; Cogstate; Eisai Inc.; Elan Pharmaceuticals, Inc.; Eli Lilly and Company; EuroImmun; F. Hoffmann-La Roche Ltd and its affiliated company Genentech, Inc.; Fujirebio; GE Healthcare; IXICO Ltd.; Janssen Alzheimer Immunotherapy Research & Development, LLC.; Johnson & Johnson Pharmaceutical Research & Development LLC.; Lumosity; Lundbeck; Merck & Co., Inc.; Meso Scale Diagnostics, LLC.; NeuroRx Research; Neurotrack Technologies; Novartis Pharmaceuticals Corporation; Pfizer Inc.; Piramal Imaging; Servier; Takeda Pharmaceutical Company; and Transition Therapeutics. The Canadian Institutes of Health Research is providing funds to support ADNI clinical sites in Canada. Private sector contributions are facilitated by the Foundation for the National Institutes of Health (www.fnih.org). The grantee organization is the Northern California Institute for Research and Education, and the study is coordinated by the Alzheimer’s Therapeutic Research Institute at the University of Southern California. ADNI data are disseminated by the Laboratory for Neuro Imaging at the University of Southern California.

## Authors’ contributions

L.M.P.S. conceptualized study design, prepared data, performed analyses, and was a major contributor in writing the manuscript. Y.K. provided feedback on analyses and contributed to the manuscript. SC and ME helped prepare figures and provided extensive feedback on manuscript preparation. E.L.A. provided guidance on interpretation of *BIN1* results and provided extensive feedback on manuscript preparation. T.J.H. performed imputation and quality control on ROSMAP genotype data. K.N. and A.J.S. provided imputed and quality-controlled ADNI genotype data. D.A.B. and J.A.S. provided ROSMAP neuropathology dataand made critical revisions to the manuscript. P.T.N. provided guidance on defining neuropathology endophenotypes and contributed to the manuscript. D.W.F. conceptualized study design and provided feedback on manuscript preparation. All authors read and approved the final manuscript.

## Acknowledgements

The NACC database is funded by NIA/NIH Grant U01 AG016976. NACC data are contributed by the NIA-funded ADRCs: P30 AG019610 (PI Eric Reiman, MD), P30 AG013846 (PI Neil Kowall, MD), P50 AG008702 (PI Scott Small, MD), P50 AG025688 (PI Allan Levey, MD, PhD), P50 AG047266 (PI Todd Golde, MD, PhD), P30 AG010133 (PI Andrew Saykin, PsyD), P50 AG005146 (PI Marilyn Albert, PhD), P50 AG005134 (PI Bradley Hyman, MD, PhD), P50 AG016574 (PI Ronald Petersen, MD, PhD), P50 AG005138 (PI Mary Sano, PhD), P30 AG008051 (PI Thomas Wisniewski, MD), P30 AG013854 (PI Robert Vassar, PhD), P30 AG008017 (PI Jeffrey Kaye, MD), P30 AG010161 (PI David Bennett, MD), P50 AG047366 (PI Victor Henderson, MD, MS), P30 AG010129 (PI Charles DeCarli, MD), P50 AG016573 (PI Frank LaFerla, PhD), P50 AG005131 (PI James Brewer, MD, PhD), P50 AG023501 (PI Bruce Miller, MD), P30 AG035982 (PI Russell Swerdlow, MD), P30 AG028383 (PI Linda Van Eldik, PhD), P30 AG053760 (PI Henry Paulson, MD, PhD), P30 AG010124 (PI John Trojanowski, MD, PhD), P50 AG005133 (PI Oscar Lopez, MD), P50 AG005142 (PI Helena Chui, MD), P30 AG012300 (PI Roger Rosenberg, MD), P30 AG049638 (PI Suzanne Craft, PhD), P50 AG005136 (PI Thomas Grabowski, MD), P50 AG033514 (PI Sanjay Asthana, MD, FRCP), P50 AG005681 (PI John Morris, MD), P50 AG047270 (PI Stephen Strittmatter, MD, PhD).

## Notes

### Author Declarations

Institutional Review Board at University of Kentucky waived ethical approval for this work.

