## Supplementary Tables S1 and S2, Figure S1 for "Genome-wide association study of multiple neuropathology endophenotypes identifies novel risk loci and provides insights into known Alzheimer’s risk loci"

### Supplementary Material

#### NACC Exclusion Criteria

National Alzheimer’s Coordinating Center (NACC) participants were excluded if they had any of the following conditions noted in the NACC Neuropathology Data Set. Variable names and descriptions are taken from <https://files.alz.washington.edu/documentation/rdd-np.pdf>. Variable descriptions may be lightly edited. Participants were not excluding for missing data in any of these fields.

| Table S1: NACC Exclusion Criteria | |
| --- | --- |
| NACC variable | description |
| NACCDOWN | Down syndrom |
| NPPDXB | Multiple system atrophy |
| NPPDXE | Malformation of cortical development |
| NPPDXD | Trinucleotide disease (Huntington disease, SCA, other) |
| NPPDXF | Metabolic/storage disorder of any type |
| NPPDXG | White matter disease, leukodystrophy |
| NPPDXH | White matter disease, multiple sclerosis or other demyelinating disease |
| NPPDXI | Contusion/traumatic brain injury of any type, acute |
| NPPDXJ | Contusion/traumatic brain injury of any type, chronic |
| NPPDXK | Neoplasm, primary |
| NPPDXL | Neoplasm, metastatic |
| NPPDXM | Infectious process of any type (encephalitis, abscess, etc.) |
| NPPDXN | Herniation, any site |
| NACCPRIO | Prion disease |
| NPPATH10 | CADASIL (hereditary stroke disorder) |
| NPALSMND | ALS/motor neuron disease (MND) |
| NPFTDTAU | FTLD with tau pathology (FTLD-tau) or other tauopathy |
| NPFTDTDP | FTLD with TDP- 43 pathology (FTLD-TDP) |
| NPOFTD | Other FTLD |
| NPPDXA | Pigment-spheroid degeneration/NBIA |

| Table S2: Participant Demographics | | | | | |
| --- | --- | --- | --- | --- | --- |
|  | NACC | ROSMAP | ACT | ADNI | Overall |
|  | (N=5625) | (N=1183) | (N=616) | (N=39) | (N=7463) |
| Sex |  |  |  |  |  |
| Female | 2809 (49.9%) | 798 (67.5%) | 345 (56.0%) | 8 (20.5%) | 3960 (53.1%) |
| Male | 2816 (50.1%) | 385 (32.5%) | 271 (44.0%) | 31 (79.5%) | 3503 (46.9%) |
| Age of Death |  |  |  |  |  |
| Mean (SD) | 81.3 (9.71) | 89.6 (6.48) | 88.4 (6.68) | 83.2 (7.90) | 83.2 (9.66) |
| Median [Min, Max] | 82.0 [39.0, 111] | 90.1 [66.0, 108] | 89.0 [70.0, 106] | 84.0 [59.0, 97.0] | 84.0 [39.0, 111] |
| APOE e4 alleles |  |  |  |  |  |
| 0 | 2533 (45.0%) | 883 (74.6%) | 443 (71.9%) | 17 (43.6%) | 3876 (51.9%) |
| 1 | 2435 (43.3%) | 280 (23.7%) | 160 (26.0%) | 17 (43.6%) | 2892 (38.8%) |
| 2 | 654 (11.6%) | 20 (1.7%) | 13 (2.1%) | 5 (12.8%) | 692 (9.3%) |
| Missing | 3 (0.1%) | 0 (0%) | 0 (0%) | 0 (0%) | 3 (0.0%) |
| CERAD |  |  |  |  |  |
| None | 519 (9.2%) | 279 (23.6%) | 139 (22.6%) | 8 (20.5%) | 945 (12.7%) |
| Mild | 467 (8.3%) | 103 (8.7%) | 159 (25.8%) | 6 (15.4%) | 735 (9.8%) |
| Moderate | 1007 (17.9%) | 399 (33.7%) | 151 (24.5%) | 2 (5.1%) | 1559 (20.9%) |
| Severe | 3627 (64.5%) | 391 (33.1%) | 167 (27.1%) | 23 (59.0%) | 4208 (56.4%) |
| Missing | 5 (0.1%) | 11 (0.9%) | 0 (0%) | 0 (0%) | 16 (0.2%) |
| Braak Stage |  |  |  |  |  |
| 0 | 71 (1.3%) | 12 (1.0%) | 20 (3.2%) | 0 (0%) | 103 (1.4%) |
| 1 | 194 (3.4%) | 75 (6.3%) | 56 (9.1%) | 2 (5.1%) | 327 (4.4%) |
| 2 | 340 (6.0%) | 109 (9.2%) | 108 (17.5%) | 7 (17.9%) | 564 (7.6%) |
| 3 | 453 (8.1%) | 281 (23.8%) | 111 (18.0%) | 3 (7.7%) | 848 (11.4%) |
| 4 | 816 (14.5%) | 370 (31.3%) | 107 (17.4%) | 1 (2.6%) | 1294 (17.3%) |
| 5 | 1418 (25.2%) | 307 (26.0%) | 129 (20.9%) | 22 (56.4%) | 1876 (25.1%) |
| 6 | 2321 (41.3%) | 18 (1.5%) | 83 (13.5%) | 4 (10.3%) | 2426 (32.5%) |
| Missing | 12 (0.2%) | 11 (0.9%) | 2 (0.3%) | 0 (0%) | 25 (0.3%) |
| Diffuse Amyloid |  |  |  |  |  |
| None | 382 (6.8%) | 222 (18.8%) | 27 (4.4%) | 1 (2.6%) | 632 (8.5%) |
| Mild | 486 (8.6%) | 351 (29.7%) | 54 (8.8%) | 3 (7.7%) | 894 (12.0%) |
| Moderate | 837 (14.9%) | 278 (23.5%) | 64 (10.4%) | 1 (2.6%) | 1180 (15.8%) |
| Severe | 2909 (51.7%) | 317 (26.8%) | 119 (19.3%) | 34 (87.2%) | 3379 (45.3%) |
| Missing | 1011 (18.0%) | 15 (1.3%) | 352 (57.1%) | 0 (0%) | 1378 (18.5%) |
| Arteriolosclerosis |  |  |  |  |  |
| None | 1173 (20.9%) | 407 (34.4%) | 7 (1.1%) | 1 (2.6%) | 1588 (21.3%) |
| Mild | 1472 (26.2%) | 398 (33.6%) | 129 (20.9%) | 24 (61.5%) | 2023 (27.1%) |
| Moderate | 1402 (24.9%) | 277 (23.4%) | 257 (41.7%) | 11 (28.2%) | 1947 (26.1%) |
| Severe | 581 (10.3%) | 81 (6.8%) | 121 (19.6%) | 3 (7.7%) | 786 (10.5%) |
| Missing | 997 (17.7%) | 20 (1.7%) | 102 (16.6%) | 0 (0%) | 1119 (15.0%) |
| Atherosclerosis |  |  |  |  |  |
| None | 1172 (20.8%) | 221 (18.7%) | 0 (0%) | 6 (15.4%) | 1399 (18.7%) |
| Mild | 1915 (34.0%) | 577 (48.8%) | 28 (4.5%) | 24 (61.5%) | 2544 (34.1%) |
| Moderate | 1421 (25.3%) | 317 (26.8%) | 166 (26.9%) | 6 (15.4%) | 1910 (25.6%) |
| Severe | 677 (12.0%) | 60 (5.1%) | 367 (59.6%) | 2 (5.1%) | 1106 (14.8%) |
| Missing | 440 (7.8%) | 8 (0.7%) | 55 (8.9%) | 1 (2.6%) | 504 (6.8%) |
| CAA |  |  |  |  |  |
| None | 1817 (32.3%) | 253 (21.4%) | 384 (62.3%) | 8 (20.5%) | 2462 (33.0%) |
| Mild | 1506 (26.8%) | 482 (40.7%) | 108 (17.5%) | 16 (41.0%) | 2112 (28.3%) |
| Moderate | 1250 (22.2%) | 261 (22.1%) | 105 (17.0%) | 8 (20.5%) | 1624 (21.8%) |
| Severe | 680 (12.1%) | 141 (11.9%) | 18 (2.9%) | 7 (17.9%) | 846 (11.3%) |
| Missing | 372 (6.6%) | 46 (3.9%) | 1 (0.2%) | 0 (0%) | 419 (5.6%) |
| LATE-NC |  |  |  |  |  |
| None | 756 (13.4%) | 509 (43.0%) | 0 (0%) | 20 (51.3%) | 1285 (17.2%) |
| Mild | 46 (0.8%) | 194 (16.4%) | 0 (0%) | 1 (2.6%) | 241 (3.2%) |
| Moderate | 259 (4.6%) | 117 (9.9%) | 0 (0%) | 14 (35.9%) | 390 (5.2%) |
| Severe | 38 (0.7%) | 267 (22.6%) | 0 (0%) | 3 (7.7%) | 308 (4.1%) |
| Missing | 4526 (80.5%) | 96 (8.1%) | 616 (100%) | 1 (2.6%) | 5239 (70.2%) |
| Lewy Body |  |  |  |  |  |
| None | 3544 (63.0%) | 861 (72.8%) | 468 (76.0%) | 17 (43.6%) | 4890 (65.5%) |
| Mild | 182 (3.2%) | 23 (1.9%) | 15 (2.4%) | 1 (2.6%) | 221 (3.0%) |
| Moderate | 666 (11.8%) | 92 (7.8%) | 49 (8.0%) | 9 (23.1%) | 816 (10.9%) |
| Severe | 701 (12.5%) | 157 (13.3%) | 51 (8.3%) | 12 (30.8%) | 921 (12.3%) |
| Missing | 532 (9.5%) | 50 (4.2%) | 33 (5.4%) | 0 (0%) | 615 (8.2%) |
| HS |  |  |  |  |  |
| Absent | 4514 (80.2%) | 1053 (89.0%) | 536 (87.0%) | 33 (84.6%) | 6136 (82.2%) |
| Present | 527 (9.4%) | 100 (8.5%) | 64 (10.4%) | 6 (15.4%) | 697 (9.3%) |
| Missing | 584 (10.4%) | 30 (2.5%) | 16 (2.6%) | 0 (0%) | 630 (8.4%) |
| Microinfarcts |  |  |  |  |  |
| Absent | 4232 (75.2%) | 820 (69.3%) | 308 (50.0%) | 31 (79.5%) | 5391 (72.2%) |
| Present | 1093 (19.4%) | 349 (29.5%) | 305 (49.5%) | 8 (20.5%) | 1755 (23.5%) |
| Missing | 300 (5.3%) | 14 (1.2%) | 3 (0.5%) | 0 (0%) | 317 (4.2%) |
| Gross infarcts |  |  |  |  |  |
| Absent | 4143 (73.7%) | 753 (63.7%) | 447 (72.6%) | 35 (89.7%) | 5378 (72.1%) |
| Present | 1095 (19.5%) | 416 (35.2%) | 169 (27.4%) | 4 (10.3%) | 1684 (22.6%) |
| Missing | 387 (6.9%) | 14 (1.2%) | 0 (0%) | 0 (0%) | 401 (5.4%) |
| Key: SD, standard deviation; Min, minimum; Max, maximum. LATE-NC, LATE neuropathologic change HS, Hippocampal sclerosisAge of Death variable is integer in NACC, ADNI, and ACT but continuous in ROSMAP. | | | | | |

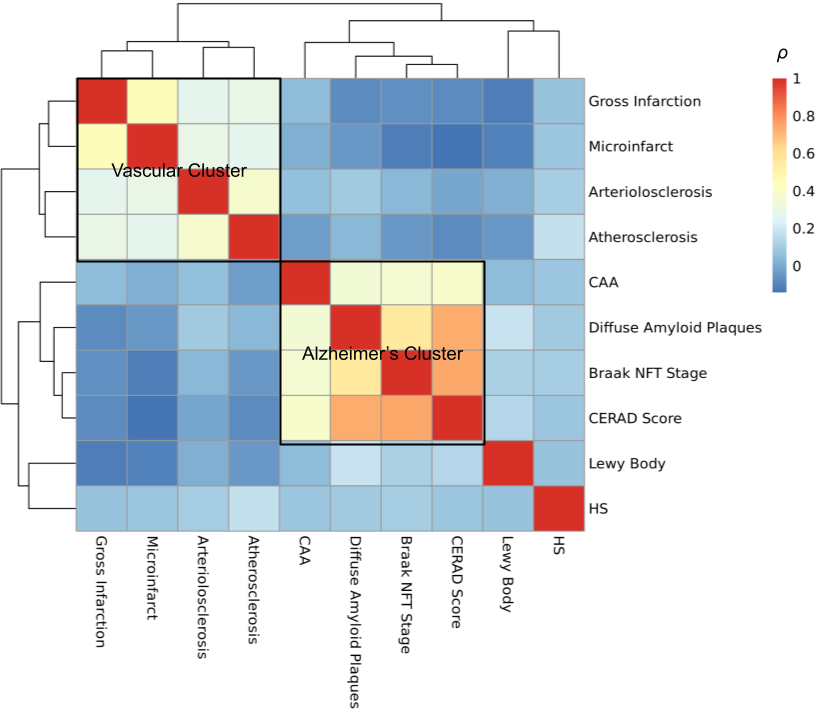
Figure S1: Dendrogram of Neuropathology Endophenotype Polychoric Correlations
